# Effect of inclusion education on the perceptions of sighted school children about the abilities of students living with severe visual impairment and blindness

**DOI:** 10.1101/2022.09.10.22279801

**Authors:** Carl Halladay Abraham, Julian Daniel Eshun, Mercy Oforiwaa Berchie, Naa Adjeley Addo

**Affiliations:** Department of Optometry and Vision Science, University of Cape Coast, Cape Coast, Ghana

## Abstract

Inclusion education was introduced to bridge the gap between access to education and individuals living with disabilities. In developing countries like Ghana, one of the primary reasons for the establishment of inclusion education was to improve the perceptions and attitudes of society from traditionally entrenched derogatory and condescending attitudes towards individuals living with disabilities, perceptions and attitudes which led to the establishment of special schools at the outskirts of towns and cities. Today, inclusion education is widely practiced and although the literature is buoyant with research on the implementation of inclusion education studies, no study has been conducted to identify the influence of inclusion education on the primary recipients, the students. Using structured interview questions exploring perceptions of sighted students in inclusion schools and sighted students in sighted only schools, we describe the differences in perceptions of these students about the abilities of individuals with visual impairment across educational prospects, leadership, participation in social activities, and routine activities. We also compared the perceptions of students living with visual impairment in inclusion schools to those in special schools. The results of the study, analyzed by Braun and Clarke’s thematic analysis showed that inclusion education has impacted and influenced positively the perceptions of the sighted students in inclusion schools about the abilities of individuals living with visual impairment compared to the perceptions held by sighted students in sighted only schools. Additionally, this study revealed that the perceptions about the abilities of students living with visual impairment in inclusion schools did not differ from those of the students living with visual impairment in special schools. This study has elucidated the need for public health education programs about the capabilities of individuals with visual impairment to foster and build healthy perceptions about them and enable a more inclusive society.

## Introduction

The evolution of inclusive education has served as a source of contention among educators and policymakers regarding its conceptualization and actualization[1]. Throughout the literature, there are varied definitions and representations of what should constitute an inclusive education[2]. The UN Convention on the Rights of People with Disabilities recognizes that persons with disabilities should have equal, unhindered rights to an education at all levels across a country in a way and manner that serves to maximize the talents of persons with disabilities, develop their intellectual and emotional capacities, as well as fully integrating them into society without discrimination delivered based on an equal opportunity[3].

Worldwide, about 15.3% of the world’s population suffers one form of disability or the other and 3% suffer severe disabilities[4]. In Ghana, according to the 2010 housing and population census, it was estimated that about 3% of Ghanaians had a disability. Among the various types of disabilities suffered in Ghana, about 40% of them were visual impairments[5]. Visual impairment is considered and defined as having visual acuities worse than 6/18 in the better-seeing eye for distance vision, and worse than N6 or 0.8M at 40cm in the better-seeing eye for near vision[6].

It has been observed that individuals with visual impairments have limited opportunities for education and employment in terms of scope and variety[7]. A direct consequence of this is poverty which gives a high correlation between visual impairment and poverty, especially in low to middle-income countries[8]. This is often facilitated by negative stereotypes that are assigned to people with visual impairments, including their ability to be productive, incompetence in managing personal affairs, at-risk for self-injury, inferiority, and socially immature[9]. Consequently, society treats persons with visual impairment as burdensome and dependent[10]. Research has shown that regardless of age, all persons with visual impairment experience a significant share of these negative societal attitudes towards them[11].

Particularly for children of school-going age with visual impairments, the situation could be notably worse for them because of the apparent manifestations and limitations of their impairment regarding mobility, interaction with peers, and participation in academic and non-academic activities. Although they might have alternative ways and unique means of going about routines to achieve comparable outcomes, they could still face those negative attitudes because their methods might be deemed out of the ‘normal’ order by their peers without disabilities. These social deficits culminate in poor peer-to-peer skills undermining their participation in school and recreational activities. Ultimately, these behaviors foster the already negative perceptions about children with disabilities and influence the set of perceptions that sighted school children have about them[12].

Traditionally, education has been organized on a two-tier basis in Ghana: the first is the special school for children with disabilities such as deafness, blindness, and mental retardation, and the second is the regular school system[13]. Ghana is a signatory to the Salamanca Statement (1994), which emphasizes more on inclusive education among member countries. As a result, the Ministry of Education, and the Ghana Education Service, in the Education Strategic plan 2003–2015, prioritized inclusive education as one of the chief policy goals[14]. Since then, children with and without disabilities have been educated together in inclusive schools in Ghana.

The implementation of inclusive education in developing countries such as Ghana is relatively at its early stages[3]. The lag in development is due to the perceptions and beliefs of people about children with disabilities and their positions in society[15]. This finding is supported by a study done by[13], who found that in Ghana, discrimination and negative attitudes still prevail against people living with disabilities. Usually, people equate the disabilities of persons with disabilities (PWDs) to inability in society[16].

Today, most students with visual impairments attend inclusive education settings[17]. However, limited studies have assessed the policy’s effects on the perceptions of the students, who are the primary recipients of the inclusive education system. Inclusive education promotes social interactions between persons with and without visual impairment. However, the extent to which such relations have impacted the traditionally held beliefs of sighted students about the abilities of students with visual impairment is unknown. In this study, we analyzed the effect of inclusion education on the perception of visually impaired and blind students by sighted students and the perceptions of visually impaired and blind students about themselves compared to what the sighted students perceive about them.

## Methods and Materials

A qualitative study design was used in this study to understand the perceptions of sighted students about students with visual impairments and the perceptions of visually impaired students about themselves compared to sighted students. A phenomenological approach was adopted to make sense of how both the visually impaired and blind students and the sighted students experience, describe, feel about, remember, and judge a particular phenomenon[18]. Thus, the phenomenological approach will allow for a detailed exploration of participants’ perceptions about the abilities of visually impaired students in inclusive education schools in Ghana.

A total of 58 participants enrolled in Junior High Schools in the Central Region randomly sampled from three schools participated in the study. The participants consisted of 10 blind students from Cape Coast School for the Deaf and Blind (special education school), 34 from Ghana National Inclusion Basic School consisting of 21 sighted students and 13 students living with blindness, and 14 sighted students from the Kwegyir Aggrey Basic School (a mainstream school for only sighted students).

Focused group discussions using simple and easy-to-understand questions around 4 broad areas: educational prospects, leadership, participation in social activities, and routine tasks to understand the perceptions of blind students in sole blind schools about themselves compared to blind students in inclusion schools and sighted students, the perceptions of sighted students in inclusion schools about blind students compared to sighted students in sole sighted schools. The participants were made to understand the importance of the study and were advised to give candid answers to the questions asked. Each focused group discussion lasted about 60 mins and was informal in nature. The interviews were recorded and transcribed verbatim. The transcripts were analyzed thematically using Braun and Clarke’s guidelines for thematic analysis[19]. Braun and Clarke’s approach to thematic analysis emphasizes identifying themes within a body of qualitative data that explores the research question. Through this process which incorporates its own methodology, useful insights are culled from the data. Braun and Clarke’s thematic analysis is very influential in qualitative thematic analysis studies mainly because it offers a clear framework for data analysis.

### Ethics

The study was approved by the Institutional Review Board of the University of Cape Coast. To conform to the Code of Ethics concerned, informed consent was obtained in writing from the parents or guardians of all the participants. Written assent was also sought from all the research participants involved, as well as consent from the schools at which the study was conducted. Confidentiality and anonymity were also ensured, as the research participants’ names and personal information were nowhere included in the research report. Participation in the study was voluntary, and the participants were made aware that they could withdraw from the research at any time. They were also informed about the likelihood of emotional trauma due to the recall of events, even though none was recorded throughout the study.

## Results

The thematic analysis yielded themes under each area of inquiry that was insightful and relevant to the study. The excerpts presented here are general representatives of similar opinions shared by other participants. In this section students with visual impairment are represented as SWVI, sighted students in inclusion schools as SSIS, and sighted students in sighted schools as SSSS.

### Educational prospects

Four themes were generated about the perceptions of SSSS and SSIS about the educational prospects of SWVI against theirs.

### Level of academic attainment

The SWVI in both the special school and inclusion school were confident about their abilities to attain any level of education they wanted, some of them implied that, like everyone else, they could dictate their way through the academic system based on their goals rather than the system dictating to them what they could do and become because of their disability,

> “*I want to get to the university. I want to attend Harvard University in America. It is a very good school, and that is where I would like to pursue my Psychology studies*.” (**CD-5)**

Generally, the SWVI from both schools anchored their ability to get to a higher (tertiary) level of education to ambition, motivation and availability of (financial) support.

> “*University. I want to learn to that height. I think the only obstacle will be the financial strength of my guardian. If he can afford it, I can learn to get to the university*.” (**CD-8)**.
>
> “*I want to go to university and even continue to do my master’s degree. Please, before I got blind, that was my aim so I don’t think that my blindness should stop me. “I can still do it if I am serious*” (**CD-3)**

Although some of them acknowledged the toll of the visual impairment on their education, none of them conceded that it is enough to truncate its length.

> “*University. I want to get there so that I can pursue my goal to become a lawyer and a presenter. It may be difficult, especially for those of us who are visually impaired, but I know that it is possible*.” (**IE (1.VI)-4**)

On the other hand, SSSS perceived that SWVI could not go beyond certain levels of academic achievements because of their impairment.

> “*Please, they can get to primary six. This is because they are blind and cannot see what will be written on the board. Again, the things that are taught at the JHS level are difficult to understand even by the sighted students; how much more the blind?*” (**KA-8**)

Whereas SSIS shared similar views with SWVI as having the capabilities to attain any level of academic achievement. They pinned the ability of the SWVI to attain any level of academic achievement as a reflection of their own motivations, readiness, and willingness and not on their disabilities.

> “*He can complete the university and even continue to pursue a master’s degree. I don’t think they have a limitation because even with their blindness over here (in this school), some are good at learning*.”). (**1.IE. S 3**).
>
> “*Masters degree or Ph*.*D*.*… there is a saying that differently-abled and disability is not inability so they can learn to the highest degree and they can even be the world best educated*.” (**2. IE. S6**).

### Academic performance and assessment

There were mixed perceptions from both the SSSS and SSIS about the ability of SWVI to top their class. They hinged their view of the inability of the SWVI to top their class on their disability, some, particularly those in the sighted schools, were skeptical about the SWVI being able to perform well in class without help from other (sighted) students or teachers and without an extended duration (relative to that of their sighted counterparts) for completion of academic exercises.

> “*They can become first in class. This is because the teachers may give them more time to answer the questions. Again, because they cannot see, the teacher could assign a student to read the questions for them so they answer and, therefore can become first*.” (**KA-2**)

It is noteworthy, however, that all the sighted students in the inclusion schools had at one point or another gone to a SWVI for an explanation on a topic or subject.

> “*Sometimes they (the visually impaired) give better explanations than some of the sighted (students), but again, it’s not in all the subjects*.” (**IE. 1S-7**)

The SWVI regarded themselves as very capable of being at the top of their class. They did not consider their disability as a stumbling block to being the best in their class.

### Participation in class activities

In a typical classroom setting, students are allowed to experience and express themselves through questioning and answering. There is an opportunity to clarify or seek clarification on a particular subject in a formal way without having to fear adverse judgment either by the teacher or colleague student. Thus, a classroom is a safe space in which a student can participate fully, mostly within the limits of formal learning.

Almost all the study participants said SWVI, like any other (sighted) student, can raise their hands in class to answer questions and that just like any other student, the teachers were appreciative of their answers.

> “*Yes (…) The teachers really listen to them and appreciate their answers and contributions a lot. Even if they give wrong answers, the teachers make good comments about their contributions and encourage them to say it again. The teachers appreciate their contributions a lot*.” (**IE. S-1**)

Some, however, had contrary views. They thought the teachers did not involve the SWVI a lot.

> “*Some of the teachers don’t call the blind people because they think that they (the blind students) don’t know the answer. “Some of the teachers don’t do well at all*.*” They don’t involve the blind people (during classes)*.” (**IE. S-2**)

Quizzes, although occasional, are a part of most academic calendars in schools. When asked if SWVI were capable of participating and representing the class or school in quiz competitions, here is what the sighted students said. To some of the SSSS, the students with visual impairment can be on the team for the quiz.

> “*Yes. They can be called. “Because they will not answer the (quiz) questions with their eyes but their mouths. So if they learn and know the answer, they can say it*” (**KA-1**)

Yet, to others, the SWVI cannot participate in quizzes at all, or could only be on the quiz team in the absence of sighted students.

> “*Please, I think that the blind cannot be chosen because, for the quiz, only those who have sight will be selected*.” (**KA-14**)
>
> “*Please, they cannot call the blind because when there is a sighted student who is equally good, the teachers will definitely choose the sighted student over the blind student for the quiz*.”.” (**KA-13**)

Similarly, the responses by the SSIS ranged from the SWVI not being able at all, they being able based on some conditions, and being very able. Here are what some had to say:

> “*Sir, please, they can’t. For us (the sighted), after the teacher teaches, we are able to take a textbook and read more about the subject but they don’t have that opportunity, so I feel we (the sighted) will always be better than them; to represent quiz competitions*.” (**IE. S-3**)
>
> “*They can and they can’t. they can do it in particular areas like English, RME, social studies, but when it come to the aspect of integrated science and mathematics, I don’t think they can because they do not cover some of the topics like ‘graphs and construction’ in mathematics*.” (**IE. 3S-5**)
>
> “*Yes. “They can call them because some of them are more intelligent than (sighted) others in the class so they can call them (the visually impaired) and even there are some questions that some of them can answer and we (the sighted) can’t so I think they can be called*.” (**IE. 3S-2**)

However, responses from most of the SWVI showed that they perceived themselves as being able to participate in quizzes, although some of them still believed their ability to do so was conditional. Notably, none of them conceded that their disability was reason enough to exclude them from participating in such quizzes;

> “*No. We haven’t participated before. I know that if we are told early to learn and prepare, we can do it, but since I came, we have not participated in any quiz before. I can’t tell of any reason because nobody has said anything (about why we haven’t been a part of the competition) before*.” (**IE (1.VI)-4**)
>
> “*No. They haven’t nominated a blind person before. I think it’s because we don’t do all the subjects (and some of the topics in the subjects we offer) and those subjects are important areas in the quiz*.” (**IE (2.VI)-2**)

Yet, some of the students with visual impairment also attributed their ability (or inability) to participate in quizzes, not to their visual impairment, but to other factors such as shyness and (un)seriousness. These two statements by two of the SWVI depict that;

“*No. But sometimes too we the visually impaired are shy about those things. I am very shy to stand in public and talk. It’s not necessarily because of the impairment but I’m shy, so I think most of my colleagues (visually impaired) too are shy*.” (**IE (1.VI)-6**)

> “*No. Sometimes, we the visually impaired we don’t involve ourselves in such things. Even if they call us to go to the center (where the quiz competition takes place), we usually prefer to stay behind and not go, so I think it’s part of the reasons why they don’t call us. We don’t show seriousness in those things*.” (**IE (1.VI)-7**)

### Appropriate education setting

Perceptions about the abilities of the SWVI to be in a particular type of education setting (either special schools or IES) and to learn different things apart from school (academic) work were expressed. Several reasons, both primary and secondary (to the impairment of the students with visual impairment), were given by the SSSS to defend why they perceived SWVI as being able to learn in special schools only. Some of the sighted students attributed their inability to their visual impairment;

> “*I prefer the blind will go to the school for the blind. This is because “over there, it will help him*.*” “Over here, they will not see (what is written on) the board and write*.” (**KA-5)**

Others also attributed their inability to secondary reasons, including inadequate knowledge by the teachers in sighted schools and inclusion schools to teach sighted and SWVI simultaneously, and impatience on the part of the teachers during teaching. For example, these are what some had to say;

> “*Please, I will prefer that they go to the school for the blind only. This is because, when they are here with us who can see, the teachers may not have the required knowledge to teach both the blind and sighted at the same time. But when he goes to the blind school, the teachers over there know everything about the blind so they will be of better help to the blind students*.” (**KA-10)**

Remarkably, almost all the SSIS perceived the SWVI as being able to, or even better off within inclusive education settings. Beyond (the students with visual impairment) being able to help themselves through, some of the sighted students said they receive help with learning from the SWVI, as in this example;

> “*I want them to be here. Sometimes, they even play with us and also they help us in learning. I don’t think that they suffer over here, so they should be here*.” (**IE. 3S-2**)

Also, some mentioned that, in addition to their ability to thrive in the IE setting, the SWVI are stimulated by the presence of the sighted students to study harder than they otherwise would;

> “*They should be here. I think that our presence challenges them to study hard, so they should be here*.” (**IE. 1S-7**)
>
> “*I feel they should be here. They learn hard when they are here. The only downside is that they make the pace of teaching and learning very slow. When the teacher teaches, he waits for them to understand before we move on to the next topic. I will prefer them to be here, nonetheless*.” (**IE (1.S)-3**)

The SWVI in the inclusion schools praised the IE setting and expressed their ability to cope with it, in addition to factors such as early completion of the curriculum and great opportunity to socialize;

> “*I prefer to be here. The teaching here is better than that at cape deaf, and we understand the things that are taught over here better. In cape deaf, sometimes it takes a week or more to finish one topic, but here, you move quite fast, and you are able to cover much*.” (**IE (1.VI)-6**)

For the SWVI in the special school, some painted IE to be an uncomfortable setting relative to their experience in the blind school. Despite its palatability, they said IE is tainted by several factors (that they have heard from their colleagues in IE school) that negate its success. Factors including teacher impatience, lack of love and unbearable pace of teaching were mentioned in these words;

> “*Please, I prefer to be here. The teachers here really teach us to understand. They are very patient with us and give us enough time to cope with the progression of learning. When there’s an example, they teach us one by one until we all understand before they move on. This may not be the case elsewhere, for example, in the inclusive school. Because they are many and several teachers take one class, the teachers rush through the teaching and illustrations, whether the blind amongst them have understood or not. Again, right after one teacher leaves, the next follows, so it makes pressure builds on the visually impaired. If the teachers at the inclusive schools even teach at our pace, I would still prefer to be here because of the special time our teachers have for us*.” (**CD-2**)
>
> “*Please, I prefer to be here. All of us here are visually impaired. So, the teachers show us so much love and give us more attention during teaching and learning. The same love could be elsewhere, but I doubt the level of attention the teachers give us here will be the same over there. Also, sometimes we hear complaints from our colleagues in the inclusive school that the teachers are not very patient with them*. (**CD-3**)

### Leadership

People with disabilities regularly contend with public attitudes towards them. Being at the forefront of the general public who assume that a disability implies functional limitations can be very daunting. In the pursuit of the objectives of this research, the area of leadership by the visually impaired was covered. Areas explored included frames of religion, politics, academia, and counsel.

### Religion

Almost all the sighted participants of the study shared similar perceptions about the abilities of people with visually impairment to become religious leaders such as pastors or Imams. To some, religious leadership is sacred, and therefore both the visually impaired and “sighted” people have equal chances at it;

> “*Yes, the visually impaired can become religious leaders. For this, it is God’s work, so the people can’t say that he is blind. “If they can do it, the people can’t say anything*” (**IE. 1.S-6**)

To the others, especially those in the sighted school (SS), mental strength is almost enough for religious leadership. Once the visually impaired has the mental preparedness to lead, he or she can do it successfully. They argued that even some of the sighted religious leaders have people who help them with their activities and read the scriptures for them to explain during religious services. So, such help will also be very pertinent to the success of the visually impaired in religious leadership. Thus, the help will in no way emphasize their disability;

> “*Please, he (the visually impaired) can do it. For example, an Imam (who is sighted) has somebody who helps them to do their things so that person (who helps) can guide him (the visually impaired) to go and lead the Muslims in prayer*” (**KA-3**)

The participants with visual impairment shared the views of the sighted participants about their (the students with visual impairment) ability to become religious leaders. In addition to their certainty about their abilities to become religious leaders, some made some stark comments about how ready they are to battle the rejections and hostilities that may come from society;

> “*I can do it. I feel no limitation at all. If the limitation comes from the church, I would ask them why my visual impairment is a problem. I can do it*.” (**CD-5**)

### Politics

According to the SSSS, people with visual impairment cannot engage in politics. Some stated that just their visual impairment precludes them from doing so, or at best can get them to the level of rallying and campaigning;

> “*Please, he can’t do it at all! Because he is blind and blind people don’t do politics. I haven’t seen one before*.” (**KA-11**)

Almost all the SSIS perceived the visually impaired as being able to engage in politics. However, they added that the visually impaired could not do so without help;

> “*Yes, I think they can. When they get the help, they can do it, but on their own, they can’t because they can’t read the document and travel alone*.” (**IE. 3S-1**)

The few who had opposing perceptions gave similar reasons like those in the SS did. It was noticed that all the participants with visual impairment perceived themselves as being able to engage in politics. However, those who expressed low to no interest in politics attributed it to lack of passion and fear of failure, other than an inability;

> “*No. Politics is something that I don’t really like. Growing up, I wished to become a president in the future, but when I understood politics more and the troubles and verbal assaults that come with failing to fulfill your promises, it has made me lose interest in it. I don’t think my visual impairment is an issue or barrier*.” (**CD-1**)

### Academic

SSSS doubted the ability of the SWVI to become student leaders for several reasons, including visual impairment, disregard by junior students, and the inability to carry out their basic duties. The following quotes evidenced these;

> “*Please, the blind cannot serve in any position. This is because, as a prefect, you a required to control the students, and the blind person can’t even see to be able to control the students to work or even keep quiet in class*.” (**KA-9**)

Findings from the SSIS about this particular subject were consistent with their general perception of SWVI in leadership. However, some were cautiously optimistic about it, stating that the overall success of the visually impaired as student leaders depends on the help of other (sighted) colleagues. However, the few who perceived otherwise gave reasons including tardy arrivals to school by most of the SWVI, other than an inability. The following remarks from some of the respondents refer.

> “*Leadership doesn’t depend on your physical status, creed, or gender. They can be if the authorities give them a chance because some of them have the potential of leadership that they can do far better than us (the sighted)*” (**IE. 2S-6**)
>
> “*They can do it. When they are in their (blind) school, they serve in these capacities, so I feel they can do the same here. The totally blind ones will just require some help or guide with movement*.” (**IE. 1S-7**)

Among the participants with visual impairment, there was a consensus about their ability to take up student leadership roles. Some backed their responses with narrations of their past or present leadership experiences. Despite their ability, others acknowledged that the visual impairment poses them some forms of difficulties in executing their duties. Yet, others contended that they had been somewhat excluded from such engagements because of ‘disabilities’ imposed on them by the school authorities. Meanwhile, others have not pursued that passion because of utter shyness. Examples of these are below;

> “*No. I haven’t. I wish to serve in a position because we are all in an inclusive school, so I want to show that what the sighted can do, we the visually impaired can also do the same. For the class prefect, the only difficulty will be to share books for your classmates because you’d have to go round the class and mention the names, and that will be impossible for me. But with the school prefect, I can*.” (**IE. 1VI-1**)
>
> “*I would want to become a school prefect. I feel they will respect the school prefect more than the class prefect. I am shy, like I said earlier, partly because of my visual impairment. I don’t want them (the sighted) to be saying that our school prefect is a blind person*.” (**IE. 1.VI-6**)

### Counsel

The sum of the responses from the SSSS showed that they were all ready to go by the counsel of SWVI. This outcome can be attributed to the fact that almost all of them focused on the counsel, rather than on the counsellor and his or her (dis)ability. The following excerpt depicts;

> “*Please, I think I will listen. “If only the advice is a good one and it will help my life I will take it even if he is blind”* (**KA-13**)

Similarly, all but one of the SSIS affirmed that they would listen to pieces of advice given to them by SWVI. Among other reasons, they pointed out that the visually impaired colleagues in the school were mostly, relatively older – a solid basis on which counsel can be sought or received;

> “*Yes. I will listen. The truth is that most of them are in the same class as us but are older than us, so they have a lot of experience, and I will listen to them*.” (**IE. 1S-4**)

For the only SSIS who posited that he would not listen to advice from the SWVI, his reasons were experience and preference. Perhaps, he has ever had bad counsel from an SWVI. But, his preference for advice from only sighted colleagues was clearly based on their visual impairment;

> “*Please, I won’t go to them for advice. Some really give bad advice. Others, too, give good ones but I would rather prefer to take advice from the sighted people. I don’t think the blind person can give me advice because they can’t see*.” (**IE. 1S-3**)

A few of the VIBS had also encountered sighted students who did not listen to their advice because of their impairment. Below is an example;

> “*Yes. But some of the sighted people we meet around feel that we are blind, so they don’t listen*.” (**CD-9**)

The rest of the SWVI in the study were confident about their ability to give counsel and be listened to. More so, some of the VIBS were put in positions that demanded the possession of some form of counselling abilities. Implicitly, the authorities deemed them fit and their counselling (leadership) abilities measurable to that of their sighted colleagues. The following excerpt refers;

> “*Yes, they listen to me. In fact, in our class, the assistant headmaster selected one other sighted person and me that if anyone has a problem, they should tell us and if we think that the problem is beyond us, we go to tell him about it so I advise a lot of people and they take it*.” (**IE. 3.VI-1**)

### Participation in social activities

Perceptions about the ability of SWVI to use social media, have hobbies and partake in entertaining activities, perform chores, and run errands were explored.

### Social media

The study participants were asked if students with visual impairment can possess and use social media accounts to interact with people. The SSSS responded that the SWVI cannot interact on social media and should not even have a social media account. In fact, some mentioned that social media is neither good nor necessary for the visually impaired. These are some examples of their words;

> “*No. Those who are blind should not have one because they are blind and cannot see even to use it*.” (**KA-1**)

To most of the SSIS, the students with visual impairment also possess the ability to have a social media account and interact with their friends. Beyond their certainty of the abilities of the visually impaired to use social media, they advocated for and had knowledge of how possible and essential some accessibility functions of smartphones aid navigation. These are what some had to say;

> “*Yes, the blind can use social media. I think the whole world should come in agreement and make special devices for blind people to use social media (more easily)*.” (**IE. 3S-5**)

The participants with visual impairment in the study had similar perceptions about their ability to use social media. Although they affirmed their ability to use it, they didn’t do so without acknowledging some difficulties. They expressed adequate knowledge about the functions of smartphones that help them to use social media applications with ease. Meanwhile, the few who were not confident about their ability to use social media did not have an idea of what smartphones or social media are. These words depict the above;

> “*Yes, a Facebook account. I had the account when I became blind, but the phone I was using didn’t have the features that will enable me to use it myself, so I was giving it to my brother to check the messages and friend requests for me. I have stopped using it now until I get a better phone, one which can talk back at us*.” (**IE. 2VI-2**)

### Hobbies and Recreational Activities

All the participants of the study perceived everyone, including the SWVI, as able to have hobbies. From the responses, almost all the participants mentioned singing, playing musical instruments, dancing, and ‘listening’ to the TV or radio as possible hobbies for children with visual impairment. Obviously, all these activities do not rely on the visual system at all or do only to a minimal extent. However, a few of the SWVI mentioned other things other than those mentioned above. For example, two of the low vision students had cycling and playing football as hobbies respectively;

> “*Mine is a football argument and “pressing phones*.*” I really love football, so everything about football is my favorite. I am partial (sighted), so I like to use the phone too. I play football a lot*.” (**IE (1.VI)-5**)

The participants with visual impairment who were totally blind also shared similar views. Besides, they highlighted other reasons for their exclusion in such activities including disability unfriendly environments;

> “*Please, I go out to just enjoy the fresh air. The kinds of games they play are not good for us (the visually impaired) because it is too physical*.” (**IE (1.VI)-2**)

### Routine tasks

Different impairments interfere with daily activities in different ways. They have different implications for individual capacity, and also generate different perceptual responses from the broader social environment. The capabilities of children with visual impairment to carry out routine tasks in school and at home were explored.

### Routine tasks in school

The everyday activities carried out by the students were sweeping the compound and the classrooms, weeding the compound, emptying the dustbin, opening the doors to the classrooms, and watering flowers and trees on the premises. The SSSS perceived the visually impaired as entirely unable to carry out these tasks. They implied that these tasks are heavily dependent on vision, the lack of which bars them from doing the chores. For example, some said;

> “*I sweep the classroom. We know that the desks are arranged, and you have to pull and push them away before you sweep so it will be impossible for blind people*.” (**KA-12**)

Some of the SSIS wanted the students with visual impairment freed from those tasks as well for the same reason-their visual impairment;

> “*I sweep the teachers’ common room. That is the work I do in my section. I don’t think that the blind can do it because it involves removing books from the table, pulling the tables aside, sweeping the room, dusting the table and rearranging the books on the table. You need your eyes to be able to do that, but the visually impaired can’t see. Maybe, the partially sighted ones can*.” (**IE. 2.VI-1**)

The SWVI had utterly different perceptions about their abilities for these tasks. Moreover, they were hypercritical about the imposed disability on them by some of the students and even some of the teachers;

> “*I usually come to school very late, so by the time I get here, they would have been done with the compound work. However, if I come to school early, I can do everything that I’m given to do because, in our place (cape deaf), we work ourselves. I can sweep, and I do sweep. There was a time I was using thread and needle to stitch my dress in class, and one asked me if we (the visually impaired) can do these things. I felt terrible about that question that day. So, I didn’t even mind the person*.” (**IE (2.VI)-2**)
>
> “*I sweep sometimes but not every day. Most times, they see us (the visually impaired) to be unable to do the compound work. Other times, they sound like they are just compassionate and would want to do the work for you. But the point is, we can do it*.” (**IE (2.VI)-3**)

### Routine tasks at home

Running errands for parents or guardians was the particular focus in this context. Most of the sighted participants of the study regarded the visually impaired as able to run errands but were not silent on its associated dangers. They implied that children with visual impairment, in the process, become acutely vulnerable to physical dangers like extortion, being cheated on by traders, and accidents. This is what one had to say;

> “*Yes. I (a sighted person) run errands for them (my parents). But I think those who are blind should not go and buy anything for their parents. “Maybe when he (the visually impaired) is going, there can be some stones or some broken bottles (in the path that can hurt them) or the person that he is going to buy the thing from will give him items that have gone bad so he can’t go. Sometimes too they can steal the money from his (the visually impaired) hands*” (**KA-3**)

However, some of the sighted participants had completely different perceptions about the subject. Some perceived the blind as unable to run errands, and therefore branded parents who make children with visual impairment run errands as wicked;

> “*Yes, they (my parents) do (make me run errands). Sir, they should not send them (the visually impaired children). The mother who will do that is wicked. If you send them (the visually impaired) and a vehicle knocks them down, what will she (the parent) say? They must only go with guiders*” (**IE. 3S-4**)

Others, on the contrary, mentioned that beyond their window of ability, running errands go a long way to help the SWVI psychologically, perhaps, making them feel more included in society. This example depicts;

> “*Yes. It is perfect for them (the visually impaired to run errands) because it will make them feel like they can also do something (that ‘normally sighted’ children can do). But they must not send them to dangerous places like the market or crowded places*. (**IE. 1S-7**)

The participants with visual impairment of the study perceived themselves as able to run errands. Although they also acknowledged some of the dangers as mentioned above, they were still confident about their ability. They pointed out that they can do far more beyond what the sighted participants perceived them to do;

> “*Yes. My mother really sends me to run errands everywhere. At times, she even signs a cheque for me to go and withdraw money from the bank. My mum always says that the fact that I am visually impaired doesn’t mean I can’t do anything. Indeed she has helped me with the self-confidence I possess now. She won’t even cook for you. She lets me cook, then she tastes and gives her comments and corrections*.” (**IE (3.VI)-1**)
>
> “*I run errands. When I’m home, I help her to knead the flour for the meat pie. It’s only the oven that she doesn’t allow me to remove the baked pastries from, but I do every other thing. She says that the intense heat will affect my eyes the more so she doesn’t allow me to go near it. Also, I sell some of the pie around my area. I go with my (sighted) brother, but we both carry baskets of pie, and he holds my hand as we go*.” (**IE (3.VI)-2**)

Additionally, some of them lamented the overblown proportions of protection by their parents or family members because of their visual impairment. Here is an example of what one said;

> “*Please, I run errands, but it’s only in the area. I don’t go outside the area because my parents fear that I may get missing or hurt myself. Meanwhile, I know that I can go (everywhere they send me to)*!” (**IE (1.VI)-3**

## Discussion

The result of this study throws light on how differently the perceptions of the abilities of visually impaired persons differed across the students by reason of their level of interactions with the SWVI.

### Educational attainment

The sum of the reasons given by the SSSS for their perceptions about the abilities of SWVI reveals perceptions that portrayed a lack of knowledge about the learning aids and teaching modalities that can help SWVI acquire the requisite knowledge to write and pass promotion examinations. On the contrary, the SSIS perceived the SWVI as being able to get to tertiary levels of education because of their knowledge of the usability of braille in notetaking and learning, and the availability of resource teachers in the IE school who provide academic support to the SWVI. Inclusive education had provided the SSIS with an experience that aided the formation of their perceptions about the abilities of SWVI regarding how farther they can go in academia. There is a need for related public education at the basic education level about the resources available to visually impaired students to help them achieve comparable academic ambitions like their sighted colleagues. The purpose of the awareness creation is not to generate sympathy and charitable feelings toward the learners with visual impairment, but to encourage an understanding of their capabilities and needs. This method has also been prescribed by Sailor et al. in Westling and Fox (2005)[30]. Some awareness activities that have been used are movies about persons with disabilities, reading books about disabilities, and workshops in the classrooms. There is the need also for the society, in general, to be sensitized to view persons with disabilities as part of the society and therefore deserve societal support by the reducing and demolishing of negative, less favorable traditionally held beliefs and practices towards persons with disabilities (PWDs).

The SWVI in the special school were found to be more comfortable in that setting. Their knowledge of some factors in the IE school, such as teacher impatience, a faster pace of teaching, and the difficulties in taking notes made them perceive the IE setting to be unfriendly and unbearable. Therefore, they expressed their inability to cope in that setting and their affinity for the special school. Based on the reports from their colleagues who were in the IE setting, the SWVIBS perceived themselves as unable to succeed in the IE setting. However, this might not be an exact and valid assertion because the SWVIIS might have exaggerated the experiences.

Jordan (2015) conducted a similar study with SWVI, who had previously attended mainstream schools[20]. He reported that those SWVI preferred to be in the special school because they indicated that their experiences in the special school are positive. Other concerns that were present in the responses he reported were lack of resources related to their visual impairment in the inclusive school setting, more friendships in the special school setting than when they attended mainstream school and a belief that their previous teachers in the mainstream school were less qualified than the teachers they currently have. Although the SWVI in Jordan’s study also reported negative experiences, it doesn’t guarantee that the SWVIBS who were in this study would also face similar situations as indicated by their colleagues in the IE setting or by the SWVI in Jordan (2015), should they be admitted into the IE school. This is because the difference between being told of an experience and living an experience could be vast and unpredictable.

### Leadership

#### Religious frame

The prevailing impression derived from the responses of all the participants was that: religion is based on the worship of a sovereign deity who is the creator of all human beings, regardless of any physical differences. Thus, religion provides a space for gregarious cultures, embracing and including everyone, as opposed to discriminating and segregating. Therefore, the opportunity to lead in such a space is opened to everyone who wishes to do so without opposition, as said in the words of this student: “*Yes, the visually impaired can become religious leaders. For this, it is God’s work, so people can’t say that he is blind. “If they can do it, the people can’t say anything*” (**IE. 1.S-6**).

In literature, Stiteler (1992) [32] identified how images of disability within Christian liturgy are most often negative, which contributes to the exclusion of people with disabilities. Also, King (1998) explored how the Black Christian church in the United States had devalued and excluded African American women with disabilities.

The above studies on religion/spirituality and disability suggest that while religion and spirituality can readily tolerate and offer positive supports to people with disabilities, their experiences within religious or spiritual communities continue to be of marginalization and exclusion. By inference, the situation could even be worse for those with disabilities who were in leadership positions in those religious settings. Nevertheless, it cannot be discounted that IE has accounted for the elevated confidence and certainty in the perceptions of the SWVIIS and the SSIS regarding the abilities of students with visual impairment to become religious leaders.

#### Political frame

The opinions of the study participants diverged concerning the potential of SWVI to engage in politics. The responses of the SSIS, SWVIIS, and SWVIBS were positive. However, they highlighted the importance of necessary aids and some structural modifications for the SWVI. Conversely, the SSSS perceived the SWVI as unable to engage in politics. Regardless of factors like competence, education, and honesty, the SSSS still did not perceive the SWVI to be worthy of leading sighted people in the arena of politics as evidenced in these words: *‘there is no way sighted voters will vote for a blind person to become their political leader*.’ The SSSS believe that visual impairment is a barrier to engagement in politics for persons with visual impairment. This is clearly stereotypical and discriminatory, yet they seemed to legitimize their claim with the fact that there has not been any visually impaired political leader in Ghana for decades[21]. Also, they mentioned that mobility challenges and the heavy reliance of politics on information transactions, many of which are documented and stored in text, require careful perusal and/or signing will further exclude the SWVI.

All the findings above are consistent with the outcome of a study conducted by Sackey[21] on the engagement of PWD in politics[21]. In Sackey’s conclusions, there was a marked correlation between the low political representation of PWDs within the district assemblies in Ghana and stigmatization against PWDs. Also, for example, after the successful election of John Mahama as the President of Ghana in December 2012, he nominated his first batch of ministers for vetting by parliament in January 2013. Dr. Henry Seidu Danaa, a lawyer with visual impairment, was one of the nominees who were assigned to the Ministry for Chieftaincy and Traditional Affairs. After his appointment, several local chiefs protested the nomination (http://thechronicle.com.gh.: 17 August 2013). They gave reasons about certain aspects of the tradition and customs in some traditional areas that did not allow PWDs to hold such high offices[21]. This incident illustrates how culture and traditions affect our perceptions.

Overall, inclusive education had some effect on the perceptions of the students in the IE school on this subject. Although the students in IE school were much more positive in their perception about the abilities of the SWVI to engage in politics, there was no bold difference between their responses and that of the students in the regular schools. Therefore, without conscious effort, just the placement of PWDs and persons without disabilities in a common school as a solution to affect such entrenched negative perceptions alone is overly simplistic, empty, and doomed to fail.

#### Academic frame

Drawing from the views the students expressed, one could infer that those pre-formed beliefs about people with visual impairment and their abilities were paramount to the formation of their perceptions regarding this theme. To address the above observation, inclusive schools need to support and implement capacity building programs for SWVI, especially those who are identified or show interest in taking up leadership roles in the school. This capacity building will better place and equip SWVI to venture and succeed in leadership in the academic environment, considering the increasing numbers of SWVI in mainstream schools. Their success will likely affect the perceptions of the sighted students about the abilities of SWVI to become leaders in school. It could also erode traditionally held beliefs and practices which devalue PWDs and completely project them as persons with deflated capabilities and potentials, unable to do anything without some form of help.

Some of the responses from the sighted students depicted that the physical and structural environment was neither disability-friendly nor accessible to the students with visual impairment as was seen in prior studies (Abraham et al., 2021; Ackah-Jnr & Danso, 2018) about similar situations in inclusion schools. Yet, they related the difficulty in mobility not to the inaccessible nature of the environment, but the individual characteristics of the SWVI. This confirms the social model of disabilities[22], which has always been on the affirmative side of the debate that the disability of the individual is not what disables them, but the social organization and architectural design of the environment within which the person lives are the sources of the disabilities. Therefore, there must be appropriate additions to the curriculum to meet the functional needs of the students with visual impairment, such as mobility and orientation around the school environment, adequate training in how to access information, independent living skills, and the use of technology and equipment. This is especially important in the early days of the students with visual impairment when they are admitted to the inclusive school.

#### Frame of Counsel

Like the perceptions about the ability of SWVI to become religious leaders, almost all the participants in the study perceived SWVI to possess the ability to give sound advice to everyone, whether sighted or visually impaired. Specifically, all the sighted participants mentioned that the SWVI are mostly relatively older than they are, although they may be classmates. This agreed with the demographic data of the participants of this study. Again, they believed that the age difference has afforded the SWVI enough time to accumulate sufficient experiences and therefore possess an exceptional body of wisdom from which they can offer counsel.

From the data collected regarding this theme, it was discovered that the inclusive school authorities had put some commendable measures in place. For instance, one of the SWVI said that “*in our class, the assistant headmaster (of the school) selected one other sighted person and me that if anyone has a problem, they should tell us and if we think that the problem is beyond us, we go to tell him about it so I advise a lot of people and they take it*.*”* Such practices are sound and could affect the perceptions of sighted students about the abilities of SWVI to give counsel. All inclusive schools could adopt this method.

### Participation in Social Activities

#### Social media

In a globalized, increasingly digitized world, the importance of computer and social media use cannot be overemphasized. Specifically, social media and technology have enhanced the development of a sense of community beyond the physical geographic location of people[23]. Social media and technology have become integral parts of our life and are increasingly becoming outlets for exercising our rights; the right to speech, the right to vote, the right to information access, and many more. As important as this is, every member of society deserves an equal chance at using social media and technology, moreover, social media and technology as tools should not promote inequality by themselves by being inaccessible to individuals living with disabilities.

The perceptions of the study participants about the abilities of SWVI to use social media were dissimilar. The SSSS perceived the SWVI as unable to use social media. In addition, they saw social media use to be unnecessary or unimportant for the SWVI because of their impairment as these two quotes from two of them depict: “*No… Those who are blind should not have one because they are blind and cannot see even to use it*.” (**SSSS-1**); and “*I don’t have an account. I think it will be difficult for the blind to use social media, and it’s not important for them*.” (**SSSS-9**). Thus, to them, visual impairment precludes a person from social media use.

Clearly, this implication reflects their pre-formed perceptions about persons with visual impairment, and ignorance about the various accessibility technologies (ATs) available in some smartphones and computers to aid users who are visually impaired. On the contrary, all the SSIS and the SWVI in the study perceived SWVI as able to use social media because they had appreciable knowledge about the ATs that aid SWVI in using social media. Moreover, they even advocated for more and upgraded forms of the ATs for SWVI. More importantly, they implied that the ATs enhance the abilities of the VIS to use social media and do not underscore their impairment. In support of this point, SWVI acknowledged the boring situation of having their messages and updates on social media read to them by sighted people because of the lack of ATs is rather over-dependent and embarrassing. This situation could make them more vulnerable to stigmatization and facilitate the formation of negative perceptions about them.

The curriculum for Information technology (IT) related subjects taught in both inclusive and non-inclusive basic schools must include contents concerning ATs and the accessibility functions that could make persons with visual impairment technologically adept[24]. This provision of knowledge could affect the perceptions of sighted students about the abilities of persons with visual impairment to use social media and computer technology in general.

#### Recreational activities

Summarizing the perceptions of the sighted students of the study on this subject, recreational activities that require eyesight and movement are mostly conceived to be untenable for people with visual impairment.

Clearly, inclusive education has not had any significant effect on the perceptions of sighted students about the abilities of SWVI to take part in routine recreational activities like football, skipping, and athletics. This observation could be because the current nature and manner of playing these games are less favorable for the SWVI. The respondents with visual impairment revealed that their participation in recreation was partly based on the availability and accessibility of certain activities. They participated in recreational activities that were within their reach, like chatting with friends during periods of recreation. The affinity for these new means of recreation for the students with visual impairment was indirectly stimulated by the barriers they face when they try to participate in the existing activities of recreation. By inference, although in an inclusive setting, the current recreational activities are acutely exclusionary, reflecting fragmentation based on visual abilities and deep disparities in social skills.

However, total exclusion from recreational activities based on one’s disability is both unfair and discriminatory. Therefore, there must be the implementation of stringent policies in the IE schools and in the society at large to ensure that persons with disabilities are not excluded from recreational activities. By this, the authorities and the students will be compelled to ensure the inclusion of the SWVI in recreational activities. For example, the Americans with Disabilities Act (ADA) forbids explicitly public and private programs from excluding individuals with disabilities from such programs solely based on their disability[25–27]. Similar acts can be adopted and adapted to the Ghanaian context to address the unique manner of exclusion persons with disabilities face, even within inclusive schools.

There are rules of adjustment and adaptation in disabled sport. Therefore, there are many recreational and sporting activities that visually impaired people could participate in. These disciplines include goal-ball, road cycling, athletics, 5-a-side football, and others[28]. Locally, some of the rules of these activities mentioned above can be adapted to modify the structure of the existing ones in the IE schools to ensure full participation by all. As a result, the potentials of SWVI in the various activities will be nurtured, and the ones who possess exceptional talents already could be identified for events like Special Olympics. In addition to enhanced self-esteem, the SWVI might also derive many health benefits from participating in physical recreation-e.g., rope jumping[29].

#### Routine Tasks

In most Ghanaian basic schools, the students have routine, assigned responsibilities to carry out either before, during, or after official school hours for teaching and learning. In an inclusive setting, that will mean the visually impaired students have a share of the responsibilities. Some of the common activities carried out daily by the students are sweeping the compound and the classrooms, weeding the compound, emptying the dustbins, opening the doors to the classrooms, and watering of flowers and trees on the premises. These activities were common in both the inclusive and mainstream settings.

The SSSS perceived the SWVI as unable to perform some of these responsibilities because the activities are visual tasks, and visual impairment completely bars the SWVI. Although some of the SSIS shared the views of the SSSS, they did not share the same reasons for their opinions. While the SSSS wanted the SWVI excluded solely because of their visual impairment, the SSIS wanted the SWVI excluded because of health risks those activities pose to SWVI and partly because of pity. For example, one SSIS said: “*Yes. The blind, too, can work. They can sweep, but we do it for them because we don’t want the dust to enter their eyes and make their eyes hurt. Also, they even force to work when we tell them not to*.” (**IE. 1S-3**). The responses from the SWVI in the study and the rest of the SSIS were contrary to that from the SSSS. They shared the common perception that the SWVI can perform those daily tasks. In addition, the SWVI in the IE school expressed their dislike for the over-protection and imposed disability on them by the sighted students and the school authorities. They implied that those restrictions from the daily activities they face detract from and retard the improvement upon their already existing competencies to do these activities.

Like the SWVIBS, the SWVIIS said that when they were in the blind school, they performed all these activities themselves. One of the SWVIIS painted the situation this way: “*I can do everything that I’m given to do because in our place (at the blind school), we work ourselves. I can sweep, and I do sweep. There was a time I was using thread and needle to stitch my dress in class (in the inclusive school), and one of the sighted students asked me if we (the visually impaired) can do these things. I felt awful about that question that day. So, I didn’t even mind the person*.” (**IE(2.VI)-2**). To buttress the point, this is what another had to say: “*When they (the sighted) see us (the visually impaired) doing something, they quickly come to take it and do it. They think we are not supposed to do those things or we can’t do them. Sometimes, they even ask us how we are able to wash our school uniforms and all that. But we do all these by ourselves because nobody does them for us. Sometimes they ask you questions like how you know when what you are washing is clean? How do you locate your washed clothes on the dry line? But we get to know all these by our sense of touch*.” (**IE(2.VI)-4**).

What the SWVI were trying to put across was that the acquisition of these competent social skills in a sighted environment is a progressive process. Therefore, the presence of the sighted students must not deny SWVI the opportunity to develop them. Again, they implied that these skills are not easily acquired, yet must be fine-tuned throughout one’s life by continuous participation in these activities. Hence, sighted students need to recognize the challenges that SWVI face when acquiring the necessary skills to enhance their social competence in a complex environment (Human 2010). Inclusive schools must ensure that SWVI are allowed to perform these activities as independently as possible. This participation with independence would enable the SWVI to develop their skills. It might as well affect the perceptions of the sighted students about the abilities of SWVI to perform these activities.

#### 4.9.2 Routine Tasks at Home

In the context of routine tasks at home, running errands for parents or guardians was the focus. The views of the sighted students and the SWVI on this theme were divergent and contrasted with each other. The SSSS perceived persons with visual impairment as unable to run errands because, in doing so, they claimed, the persons with visual impairment become vulnerable to dangers like being knocked down by a vehicle, stumbling and falling, and extortion. In fact, the SSSS went on to brand parents who send their children with visual impairment on errands to be wicked as depicted in these words: “*They should not send them (the visually impaired children). The mother who will do that is wicked. If you send them (the visually impaired) and a vehicle knocks them down, what will she (the parent) say?*” (**IE. 3S-4**).

Some of the SSIS were pitiful of the SWVI and therefore mentioned that they should be made to participate in only the tasks that are in the immediate surroundings of the SWVI, confined within the walls of their residence. Yet, others, on the contrary, mentioned that beyond deepening the depths of their abilities, running errands go a long way to help the SWVI psychologically and, perhaps, make them feel more included in the society. In support of this point made by the sighted student, this is what one of the SWVI had to say: “*Yes. My mother really sends me to run errands everywhere. At times, she even signs a cheque for me to go and withdraw money from the bank. My mum always says that the fact that I am visually impaired doesn’t mean I can’t do anything. Indeed she has really helped me with the self-confidence I possess now. She won’t even cook for you. She lets me cook, then she tastes and gives her comments and corrections*.” (**IE(3.VI)-1**).

The SWVI implied that they could do more than what the sighted participants perceive them to be able to do. Although they acknowledged some of the dangers expressed by the sighted students, they were still confident about their ability to run errands. Additionally, some of them lamented the overblown proportions of protection by their parents or family members because of their visual impairment. This is an example of what one said; “*Please, I run errands, but it’s only in the area. I don’t go outside the area because my parents fear that I may get missing or hurt myself. Meanwhile, I know that I can go (everywhere they send me to)*!” (**IE(1.VI)-3**). The SWVI implied that these skills are essential to develop now, the lack of which will retard or prevent them from attaining independence later in life. Therefore, they believed that, despite their impairment, they should not be excluded from some routine activities at home, like running errands. In the view of the SWVI, “it is the society which disables physically impaired people. Disability is something imposed on top of their impairments by the way they are unnecessarily isolated and excluded from full participation in society.” (Shakespeare & Watson, 2002:3).

## Conclusion

It is noteworthy that IE has affected positively the perceptions of the SSIS regarding the academic achievement of SWVI. Therefore, if IE is better strengthened and implemented, it is likely to help to integrate PWVI into society better and create more opportunities for them. Also, the barriers and challenges they face because of negative perceptual dispositions of sighted persons are likely to diminish.

## Data Availability

Data will be made available on request. Requests for data should be sent to the corresponding author.

## REFERENCES

1. Berlach RG, Chambers DJ. Inclusivity imperatives and the Australian national curriculum. Educ Forum. 2011;75: 52–65. doi:10.1080/00131725.2010.528550

2. Forlin C, Chambers D, Loreman T, Deppler J, Sharma U. Inclusive education for students with disability: A review of the best evidence in relation to theory and practice. Educ Pap J Artic. 2013 [cited 16 Aug 2022]. Available: https://researchonline.nd.edu.au/edu_article/141

3. Convention on the Rights of Persons with Disabilities – Articles | United Nations Enable. [cited 16 Aug 2022]. Available: https://www.un.org/development/desa/disabilities/convention-on-the-rights-of-persons-with-disabilities/convention-on-the-rights-of-persons-with-disabilities-2.html

4. The global burden of disease : 2004 update. [cited 16 Aug 2022]. Available: https://apps.who.int/iris/handle/10665/43942

5. Ghana - Population and Housing Census 2010 - Ghana. [cited 16 Aug 2022]. Available: https://www2.statsghana.gov.gh/nada/index.php/catalog/51

6. Kv V, Vijayalakshmi P. Understanding definitions of visual impairment and functional vision. Community Eye Heal. 2020;33: S16. Available:/pmc/articles/PMC8115704/

7. Blackorby J, Wagner M. Longitudinal Postschool Outcomes of Youth with Disabilities: Findings from the National Longitudinal Transition Study: http://dx.doi.org/101177/001440299606200502. 2016;62: 399–413. doi:10.1177/001440299606200502

8. Holden BA. Blindness and poverty: a tragic combination. Clin Exp Optom. 2007;90:401–403. doi:10.1111/J.1444-0938.2007.00217.X

9. Rusu Mocănașu D. STEREOTYPES ABOUT BLINDNESS AND PEOPLE WITH VISUAL IMPAIRMNTS. Int Multidiscip Sci Conf Dialogue between Sci Arts, Relig Educ. 2019;3: 234–241. doi:10.26520/MCDSARE.2019.3.234-241

10. Hazzard A. Children’s Experience With, Knowledge of, and Attitude Toward Disabled Persons: http://dx.doi.org/101177/002246698301700204. 2016;17: 131–139. doi:10.1177/002246698301700204

11. Fraser S, Beeman I, Southall K, Wittich W. Stereotyping as a barrier to the social participation of older adults with low vision: a qualitative focus group study. BMJ Open. 2019;9: e029940. doi:10.1136/BMJOPEN-2019-029940

12. Babik I, Gardner ES. Factors Affecting the Perception of Disability: A Developmental Perspective. Front Psychol. 2021;12. doi:10.3389/FPSYG.2021.702166

13. Agbenyega J. Examining Teachers’ Concerns and Attitudes to Inclusive Education in Ghana. Int J Whole Sch. 2007;3: 41–56.

14. Mantey EE. Discrimination against children with disabilities in mainstream schools in Southern Ghana: Challenges and perspectives from stakeholders. Int J Educ Dev. 2017;54: 18–25. doi:10.1016/j.ijedudev.2017.02.001

15. Anthony J. Conceptualising disability in Ghana: implications for EFA and inclusive education. https://doi.org/101080/136031162011555062. 2011;15: 1073–1086. doi:10.1080/13603116.2011.555062

16. Avoke M. Models of Disability in the Labelling and Attitudinal Discourse in Ghana. http://dx.doi.org/101080/0968759022000039064. 2010;17: 769–777. doi:10.1080/0968759022000039064

17. Ametepee LK, Anastasiou D. Special and inclusive education in Ghana: Status and progress, challenges and implications. Int J Educ Dev. 2015;41: 143–152. Available: https://www.academia.edu/11449098/Special_and_inclusive_education_in_Ghana_Status_and_progress_challenges_and_implications

18. Patton MQ. What Brain Sciences Reveal About Integrating Theory and Practice: http://dx.doi.org/101177/1098214013503700. 2013;35: 237–244. doi:10.1177/1098214013503700

19. Braun V, Clarke V. Using thematic analysis in psychology. Qual Res Psychol. 2006;3: 77–101. doi:10.1191/1478088706QP063OA

20. Jordan J. Satisfaction of students with visual impairment within different school settings. Educ Spec 2009-2019. 2015 [cited 19 Aug 2022]. Available: https://commons.lib.jmu.edu/edspec201019/10

21. Sackey E. Disability and political participation in Ghana: an alternative perspective. Scand J Disabil Res. 2015;17: 366–381. doi:10.1080/15017419.2014.941925/METRICS/

22. Oliver M. The social model of disability: thirty years on. https://doi.org/101080/096875992013818773. 2013;28: 1024–1026. doi:10.1080/09687599.2013.818773

23. Drahošová M, Balco P. The analysis of advantages and disadvantages of use of social media in European Union. Procedia Comput Sci. 2017;109: 1005–1009. doi:10.1016/j.procs.2017.05.446

24. Abraham CH, Boadi-Kusi B, Morny EKA, Agyekum P. Smartphone usage among people living with severe visual impairment and blindness. Assist Technol. 2021 [cited ?8 Sep 2022]. doi:10.1080/10400435.2021.1907485

25. Appenzeller H, Appenzeller H. Risk management in sport : issues and strategies. 2012; 529.

26. Block ME. Americans with Disabilities Act: Its Impact on Youth Sports. J Phys Educ Recreat Danc. 1995;66: 28–32.

27. Bourne RRA, Steinmetz JD, Flaxman S, Briant PS, Taylor HR, Resnikoff S, et al. Trends in prevalence of blindness and distance and near vision impairment over 30 years: An analysis for the Global Burden of Disease Study. Lancet Glob Heal. 2021;9: e130–e143. doi:10.1016/S2214-109X(20)30425-3

28. Kamelska AM, Mazurek K. The Assessment of the Quality of Life in Visually Impaired People with Different Level of Physical Activity. Phys Cult Sport Stud Res. 2015;67: 31–41. doi:10.1515/PCSSR-2015-0001

29. Chen CC, Lin SY. The impact of rope jumping exercise on physical fitness of visually impaired students. Res Dev Disabil. 2011;32: 25–29. doi:10.1016/J.RIDD.2010.08.010

30. Westling, D.L & Fox, L. (2005). Teaching Students with Severe Disabilities. Upper Saddle River, New Jersey: Prentice-Hall Inc.

31. Jordan, J. (2015). Satisfaction of students with visual impairment within different school settings.

32. Stiteler, V.C.J. (1992). ‘Singing Without a Voice: Using Disability Images in the Language of Public Worship.’ Liturgical Ministry, 1, pp. 140–142.

